# Applying a Prospective Space-Time Scan Statistic to Examine the Evolution of COVID-19 Clusters in the State of São Paulo, Brazil

**DOI:** 10.1101/2020.06.04.20122770

**Authors:** R.V. Ferreira, M.R. Martines, R.H. Toppa, L.M. Assunção, M.R. Desjardins, E.M. Delmelle

## Abstract

We present the first geographic study that uses space-time statistics to monitor COVID-19 in Brazil. The first cases of COVID-19 were confirmed in December 2019 in Wuhan, China, caused by the contamination of the SARS-CoV-2 virus, and quickly turned into a pandemic. In Brazil, the first case occurred on January 23rd, 2020 but was officially reported by the Brazilian Ministry of Health on February 25th. Since then, the number of deaths and people infected by COVID-19 in Brazil have been steadily increasing. Despite the underreporting of coronavirus cases by government agencies across the country, the State of São Paulo has the highest rate among all Brazilian States. Thus, it is essential to detect which areas contain the highest concentration of COVID-19 to implement public policies, to mitigate the spread of the epidemic. To identify these critical areas, we utilized daily confirmed case data from the Brasil.IO website between February 25^th^, 2020 to May 5^th^, 2020; which were aggregated to the municipality level. A prospective space-time scan statistic was applied to evaluate possible active clusters in three different time periods. The results visualize the space-time evolution and dynamics of COVID-19 clusters in the State of São Paulo. Since the first study period, the results highlight approximately 4.6 times the number of municipalities belonging to a significant cluster with a RR>1 on May 5^th^. These results can inform health authorities and public management to take the necessary measures to minimize the transmission of COVID-19 and track the evolution of significant space-time clusters.

**HIGHLIGHTS:**

- Prospective space-time statistics can improve COVID-19 surveillance in Brazil
- All statistically significant clusters are located near São Paulo Municipality
- There are municipalities with relative risk highest than one in the countryside
- 4.6 times the number of municipalities belong to a significant cluster on May 5th

## Introduction

After five months of the novel coronavirus disease’s (COVID-19) first identification in Wuhan city (Hubei province, China) in December of 2019, a new research reports the virus was already spreading late December 2019 in Paris, in a patient that not visited Wuhan, indicating that the epidemic started earlier (Deslandes, et al. 2020). The World Health Organization (WHO) declared COVID-19 as a pandemic on March 11th, 2020 (WHO, 2020). As of May 14th, 2020, there are over 4.3 million confirmed cases and almost 300,000 deaths, globally (Dong et al. 2020). The asymptomatic rate of may be between 40%-60% (He et al. 2020; Nishiura et al. 2020), essentially indicating that a large proportion of infected individuals are “silent carriers”. Furthermore, the basic reproductive number (R_0_) is estimated between 1.4 and 6.49, making the SARS-CoV-2 virus substantially more contagious than influenza and other coronaviruses like MERS-CoV and SARS-CoV (Liu et al. 2020). Symptomatic cases are mostly mild (approximately 80%), while severe cases include clinical manifestations such as pneumonia, multi-organ failure, and death (Ruan et al. 2020; Mahase 2020).

The highest proportion of deaths occur in the elderly, immunocompromised populations, and those with preexisting conditions such as heart disease and hypertension (Wu and McGoogan 2020). Conversely, severe outcomes and deaths have also been reported in children, (Deyà-Martínez et al. 2020; Fabre et al. 2020; Jones et al. 2020). Furthermore, there is currently no known level of immunity to SARS-CoV-2 (Osman et al. 2020), therefore, spatiotemporal surveillance of COVID-19 can be a powerful tool to mitigate transmission risk (Yang et al. 2020). In this study, we focus our efforts on the State of São Paulo, Brazil.

Brazil reported the first case on January 23rd 2020, in São Paulo Municipality, but the precise time in which the virus began to spread locally is currently unknown, but probably started to spread locally before the date set by the government (Delatorre et. al. 2020). However, the Brazilian Ministry of Health reported officially that the first case occurred on February 25th, 2020, in São Paulo State (Brasil, 2020a). The World Health Organization (WHO) declared COVID-19 a pandemic on March 11th, 2020 (WHO, 2020). In Brazil on May 11th, 2020, there were 162,622 confirmed cases, 11,123 confirmed deaths, and a lethality rate of 6.8% (Brasil, 2020b). However, a high underreporting of deaths is expected due to testing lags and lack of testing resources and utilization (Alonso et al. 2020). Even more critical is that Brazil exhibits one of the highest transmission rate among the 48 countries surveyed by the Imperial College of London (BHATIA, 2020). Large cities such as São Paulo and Rio de Janeiro are emerging as the main epicenters of the epidemic (The Lancet Editorial, 2020). There are indications of faster spreading within the State of São Paulo (FIOCRUZ, 2020), where smaller population municipalities have limited access to hospital care.

Monitoring the spread of COVID-19 is essential to develop health strategies, such as making tests and hospital beds available. The prospective space-time scan statistic (Kulldorff, 1997) is a powerful exploratory approach for detecting active and emerging clusters of disease, identifying locations that are currently experiencing excess incidence of cases on the most recent time period in a dataset. The statistic determines if the observed space-time patterns of disease are randomly distributed or exhibit statistically significant clustering. It utilizes cylindrical scanning windows of different spatial and temporal dimensions to systematically scan the study area and period for excess incidence of disease cases (e.g. more observed than expected cases given baseline conditions). The prospective version of the scan statistic will disregard clusters that may have previously existed and only detect clusters that are an active and emerging public health threat (Kuldorff 2001). This is different than the retrospective space-time scan statistic, which detects clusters at any point during the study period, regardless when the clusters “disappear” (Desjardins et al. 2018; Owusu et al. 2019; Whiteman et al. 2019).

Regarding COVID-19 surveillance, the prospective space-time scan statistic has been applied to daily case data at the municipality level in the United States, analyzing a 65-day period as well as active and emerging clusters (Desjardins et al., 2020). This approach could inform decision makers to improve resource allocation and justify continued social distancing and stay-at-home orders. It is essential to clarify that this type of study can be repeated when more data is available, as to support the COVID-19 surveillance (Desjardins et al., 2020). In Brazil, as the State Health Secretariats (SHS) are updating the data daily and making them public, researchers will be able to locate new emerging clusters, as well as indicate the areas in which the COVID-19’s transmission has decreased.

Regarding COVID-19 surveillance efforts in Brazil, the Ministry of Health reports daily confirmed cases and deaths; while also developing a COVID-19 app and WhatsApp channel (de Oliveira et al. 2020). Tarrataca et al. (2020) applied a susceptible, exposed, infected, removed (SEIR) model and developed several lockdown scenarios and lack thereof. Another study in Brazil estimated the number of underreported cases and deaths (Riberio and Bernardes 2020). However, there is currently a lack of spatially explicit studies in Brazil, especially space-time cluster detection of the known burden of COVID-19 across the country at the municipality level.

In this research we by the abovementioned app by analyzing the State of São Paulo, the most affected by COVID-19 in Brazil, that registered 43,131 cases (25.6%) of the 168,331 on May 12th, 2020, (Brasil, 2020b). Utilizing a prospective space-time scan statistic, our objective was to detect new emerging clusters of COVID-19 in the State of São Paulo’s municipalities, considering three periods of analysis: (1) February 25th – March 24nth, 2020; (2) February 25th – April 15th, 2020; and (3) February 25th – May 5th, 2020. Examining these three periods, we intend to demonstrate the evolution of the relative risk in different regions and municipalities in the State of São Paulo.

## Data and Methods

On March 1st, 2020, the Health Surveillance Secretary of the Ministry of Health of Brazil stated that a COVID-19 case is defined as confirmed if tested in a laboratory with a positive result in real-time with Reverse-Transcriptase Polymerase Chain Reaction (RT-PCR) test, following the protocol of the University Charité (Berlin, Germany). Real time RT-PCR is a nuclear-derived method for detecting the presence of specific genetic material from any pathogen, including a virus – scientists can see the results almost immediately while the process is still ongoing; conventional RT-PCR only provides results at the end. In Brazil, the notification system is decentralized, and the municipalities can proceed with tests in private or public laboratories. Then, the results are notified to the State Health System, which in turn notifies the national Government.

The State of São Paulo has approximately 45.9 million inhabitants and an area of 95.8 square miles. On March 24th, the Governor of São Paulo decreed social distancing to prevent the spread of the novel coronavirus. As the incubation period for the novel coronavirus disease (COVID-19) is well supported by evidence to be around 14 days (Lauer, et. al. 2020), we selected three separate space-time analyses: from February 25th to March 24th, from February 25th to April 15th, and from February 25th to May 5th, 2020 – which includes five incubation periods of onset of the most current COVID-19 case in the dataset (Figure 1).

**Figure 1.**
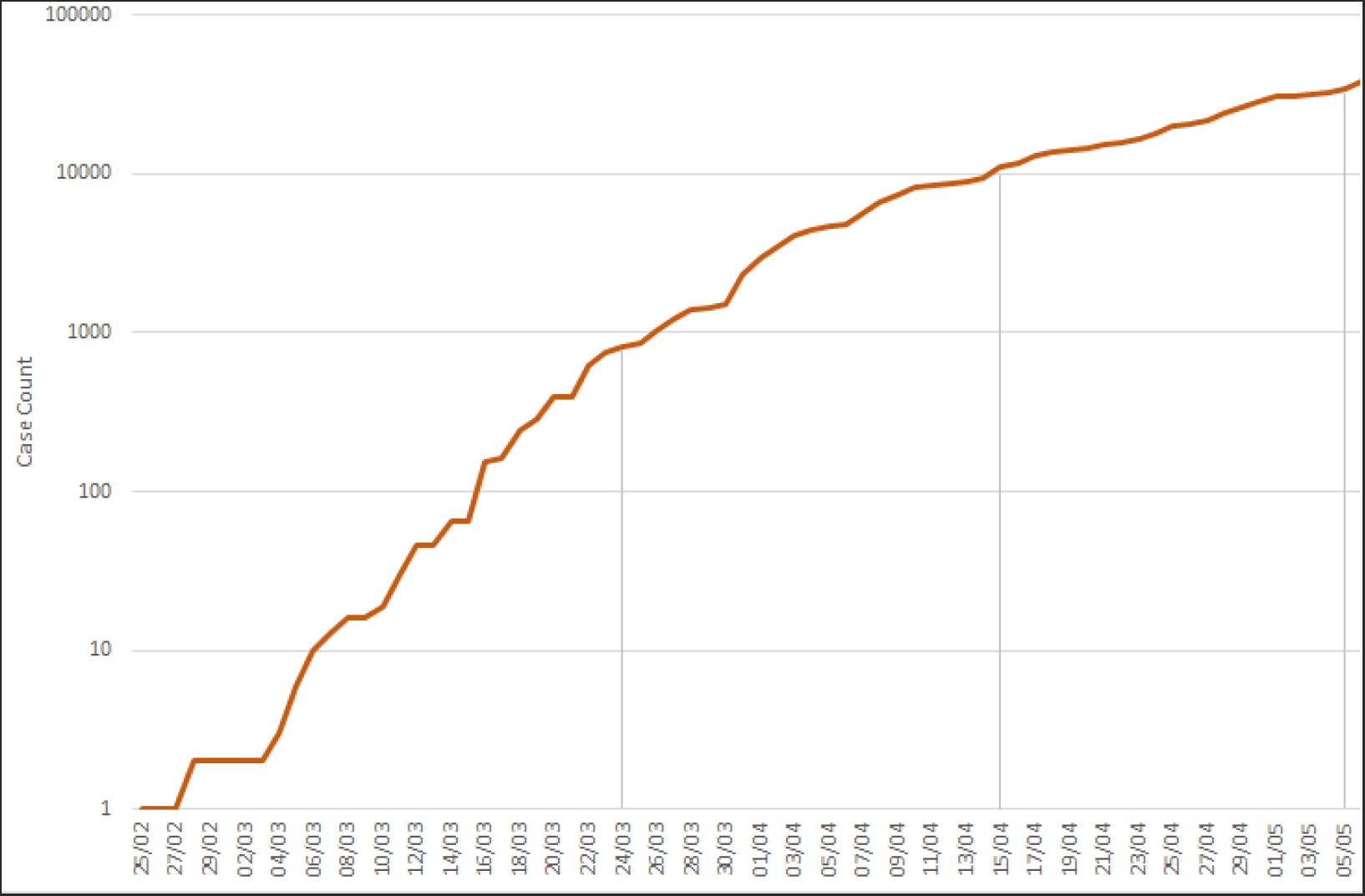
Logarithmic graph of cumulative number of COVID-19 cases in the State of São Paulo between February 25th and May 5th, 2020 (used for the statistical analysis)

We utilized confirmed daily case data at the municipality level in the State Health Secretary of São Paulo’s freely available on its official page (São Paulo, 2020a). The data contains information on 645 municipalities of the State of São Paulo and is updated daily, although we only use records between February 25th, 2020 and May 5th, 2020. We used a Geographic Information System to geocode the locations of COVID-19 cases and aggregated them to the available municipalities. The municipality-level GIS files were retrieved from the Instituto Brasileiro de Geografia e Estatística (IBGE); English – The Brazilian Institute of Geography and Statistics.

The detection of active clusters was performed using the Poisson space-time scan statistic method (Kuldorff, 2001). Analyses were done using SaTScan, a free software developed for cluster detection. The prospective space-time statistic detects active clusters of disease, i.e. excess incidence of cases during the last time period of analysis (Jones et. al, 2006). The statistic systematically implements moving cylinders to scan the study area. The cylinders are centered on the centroid of the municipalities, the base of the cylinder is the spatial scanning window, and the height represents the temporal scanning window. The cylinders are expanded until a maximum spatial and temporal upper bound is reached. We defined the upper bounds to have a maximum spatial and temporal scanning window size of 10% of the population at-risk to avoid extremely large clusters; and 50% of the study period, respectively. Each cluster’s duration was set to a minimum of 2 days and a cluster must contain a minimum of five confirmed cases of COVID-19 (Desjardins et al. 2020).

We used a prospective Poisson model to detect space-time clusters that are still occurring or active on the last day of analysis (Kulldorff, 2001; Kulldorff et al., 1998; Desjardins et al. 2020). We assume that the COVID-19 cases in our study area follow a Poisson distribution. While the null hypothesis states that the model reflects a constant risk (i.e. no anomalous clustering of COVID-19). The alternative hypothesis states that the number of observed cases exceed the number of expected cases derived from the null model. The expected cases are calculated by multiplying the population in the cylinder by the total COVID-19 rate in the cylinder (i.e. p * C/p). We assume that the population is static for time period.

Next, a maximum likelihood ratio test identifies cylinders with an elevated risk for COVID-19. The cylinder has an elevated risk when the likelihood ratio is greater than one; where the rate of cases in the cylinder is greater than the rate outside of the cylinder. To derive statistical significance, 999 Monte Carlo simulations are computed, while we report clusters at the 95% confidence level (i.e. p < 0.05). Therefore, 999 likelihood ratios are computed for each cylinder, where each are a potential cluster.

To circumvent the issue of assuming the municipalities belonging to a cluster are homogenous, we also report and map the relative risk for each municipality in each study period. The relative risk is defined the estimated risk of COVID-19 within a municipality divided by the risk outside of the municipality (i.e. everywhere else). As the pandemic continues, new data can be added the prospective space-time scan statistic to monitor active clusters and identify areas that no longer are experiencing excess incidence based on available confirmed cases (i.e. areas that no longer have an excess public health risk).

## Results

### First period of analysis – February 25th – March 24th, 2020

We did not detect any statistically significant emerging space-time clusters of COVID-19 between the first period of analysis – 28 days, two incubation periods – (Figure 2A). However, we observed five municipalities with a RR>1 outside the cluster for this first period, that is, with more observed than expected cases (Table 1). *São Paulo* Municipality has the highest relative risk (RR = 28.03), followed by *São Caetano do Sul* (RR = 3.42), *Jaguariúna* (RR = 2.38*), Santana de Parnaíba* (RR = 1.97) and *Cotia* (RR = 1.1).

**Table 1.** Municipalities of São Paulo State, Brazil, with relative risk (RR) > 1, observed/expected cases and population, within the time period between February 25th and March 24th, 2020.

| *Municipality* | *Population* | *Observed cases* | *Expected cases* | *RR* |
| --- | --- | --- | --- | --- |
| São Paulo | 12,252,023 | 306 | 89.65 | 28.03 |
| São Caetano do Sul | 161,127 | 4 | 1.18 | 3.42 |
| Jaguariúna | 57,488 | 1 | 0.42 | 2.38 |
| Santana de Parnaíba | 139,447 | 2 | 1.02 | 1.97 |
| Cotia | 249,210 | 2 | 1.82 | 1.1 |

### Second period of analysis – February 25th – April 15th, 2020

We detected seven statistically significant emerging space-time clusters of COVID-19 in São Paulo State for this second period of analysis (50 days, approximately three incubation periods), which included 353 municipalities. In this second period there were two observed clusters with a RR>1 (cluster 1 – RR = 3.29; and cluster 3 – RR = 2.64), including 11 and 17 municipalities, respectively (Table 2). Both are located in the East region of São Paulo State, near São Paulo Municipality (Figure 2B).

**Table 2.** Emerging space-time clusters of COVID-19 from February 25th – April 15th, 2020 show the relative risks (RR) for the clusters identified in the São Paulo State, Brazil.

| *Cluster* | *Duration* | *P* | *Observed cases* | *Expected cases* | *RR* | *# of municipalities* | *Population* | *# of municipalities with RR>1* |
| --- | --- | --- | --- | --- | --- | --- | --- | --- |
| 1 | 3/30-4/15 | <0.001 | 943 | 305.05 | 3.3 | 11 | 3,845,800 | 4 |
| 2 | 3/22-4/15 | <0.001 | 118 | 522.04 | 0.2 | 203 | 4,475,331 | 2 |
| 3 | 3/31-4/15 | <0.001 | 641 | 252.45 | 2.6 | 17 | 3,381,536 | 6 |
| 4 | 3/22-4/15 | <0.001 | 86 | 421.67 | 0.2 | 61 | 3,614,906 | 0 |
| 5 | 3/22-4/15 | <0.001 | 22 | 168.99 | 0.1 | 33 | 1,448,732 | 0 |
| 6 | 3/22-4/15 | <0.001 | 36 | 127.66 | 0.3 | 9 | 1,094,410 | 0 |
| 7 | 3/22-4/15 | <0.001 | 21 | 96.37 | 0.2 | 19 | 826,174 | 0 |

**Figure 2.**
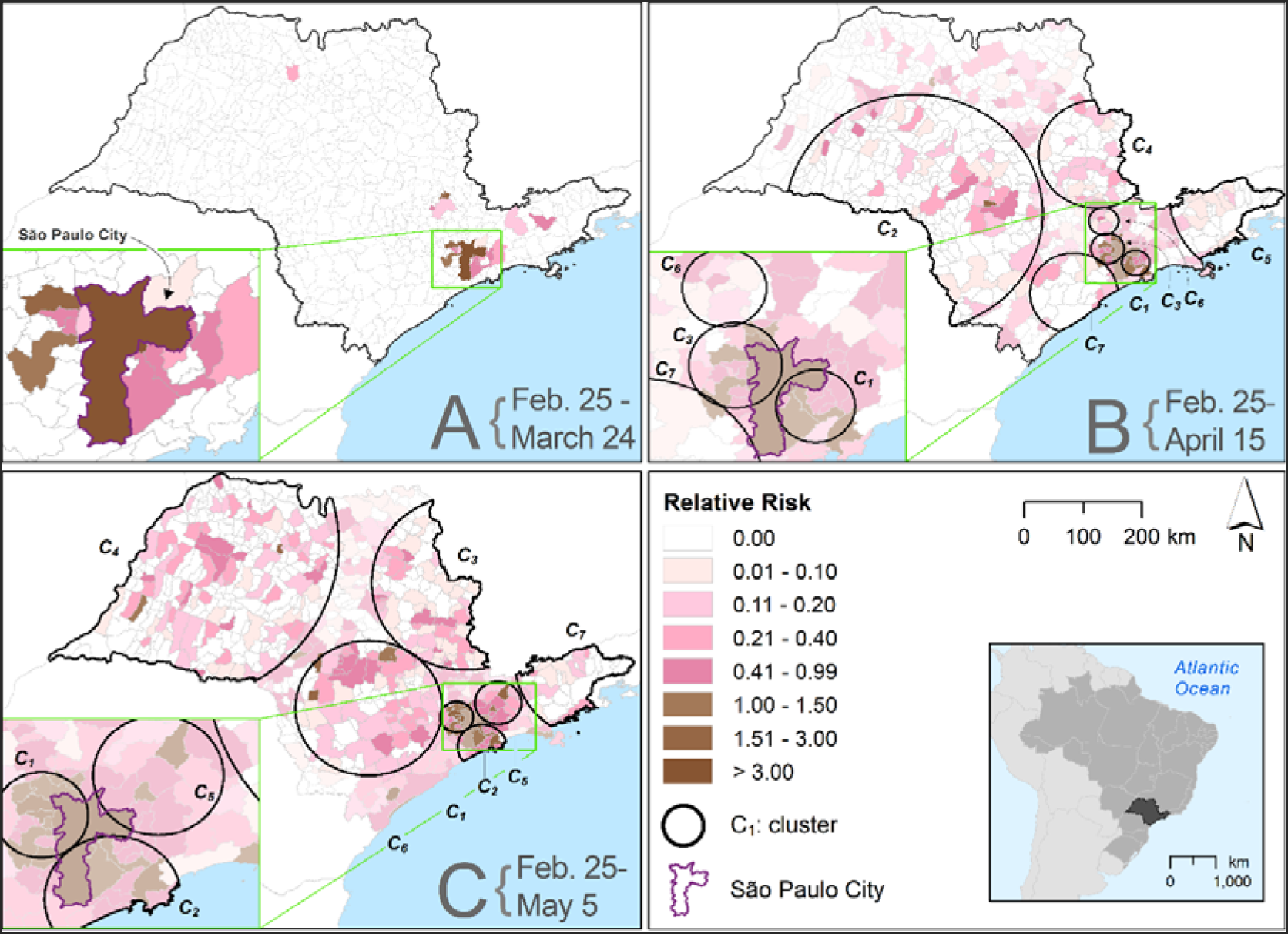
Spatial distribution of emerging space-time clusters of COVID-19 for the municipalities of São Paulo State, Brazil. A – First period of analysis – February 25th – March 24th, 2020 (28 days). B – Second period of analysis – February 25th – April 15th, 2020 (50 days; C1 – RR = 3.29; C3 = 2.64; the other clusters did not show a RR>1). C – Third period of analysis – February 25th – May 5th, 2020 (70 days; C1 – RR = 3.71; C2 – RR = 3.12; C5 – RR = 2.68; the other clusters did not show a RR>1).

Cluster 1 has 943 observed cases, and *São Caetano do Sul* Municipality has the highest relative risk (RR = 2.23) with 85 observed cases for 38.34 expected cases, followed by *Santos* Municipality (RR = 2.12 with 216 observed cases for 103.11 expected cases). *São Bernardo do Campo* Municipality is also in cluster 1 (RR = 1.16), and *Santo André* Municipality (RR = 1.09) with RR>1. Cluster 3 contains 17 municipalities including six municipalities with a RR>1 and 641 observed cases (Table 2). *Caieiras* Municipality has the highest relative risk (RR = 1.74), followed by *Taboão da Serra* (RR = 1.34), *Santana de Parnaíba* (RR = 1.24), *Cotia* (RR = 1.18), *Franco da Rocha* (RR = 1.12), and *Osasco* (RR = 1.04).

### Third period of analysis – February 25th – May 5th, 2020

We detected seven statistically significant emerging space-time clusters of COVID-19, in our third period of our analysis (70 days, five incubation periods), covering 513 municipalities of the *São Paulo* State. Clusters 1, 2, and 5 show the highest relative risks (Table 3) with a RR = 3.71, RR = 3.12, and RR = 2.68, respectively. All of them are located near *São Paulo* Municipality, in the East region of the State (Figure 2C).

**Table 3.** Emerging space-time clusters of COVID-19 from February 25th – May 5th, 2020 show the relative risk (RR) for the clusters detected in São Paulo State, Brazil.

| Cluster | Duration | P | Observed cases | Expected cases | RR | # of municipalities | Population | # of municipalities with $RR > 1$ |
| --- | --- | --- | --- | --- | --- | --- | --- | --- |
| 1 | 4/15-5/5 | <0.001 | 2,598 | 741.98 | 3.7 | 17 | 3,381,536 | 11 |
| 2 | 5/15-5/5 | <0.001 | 2,899 | 985.48 | 3.1 | 14 | 4,491,304 | 5 |
| 3 | 5/1-5/5 | <0.001 | 534 | 1,641.67 | 0.3 | 87 | 4,489,123 | 0 |
| 4 | 4-1-5/5 | <0.001 | 583 | 1,660.57 | 0.3 | 263 | 4,540,784 | 2 |
| 5 | 4-24-5/5 | <0.001 | 1,118 | 425.91 | 2.7 | 15 | 3,396,868 | 2 |
| 6 | 4-1-5/5 | <0.001 | 758 | 1,635.26 | 0.4 | 85 | 4,471,575 | 3 |
| 7 | 4/1-5/5 | <0.001 | 108 | 485.36 | 0.2 | 32 | 1,327,200 | 0 |

Cluster 1 includes 11 municipalities with a RR>1 and the highest relative risk observed is *Barueri* Municipality (RR = 2.03). Cluster 2 has five municipalities with a RR>1 and *São Caetano do Sul* Municipality has the highest relative risk (RR = 1.84). Cluster 5 has two municipalities with a RR>1 and *Igaratá* Municipality is the highest relative risk (RR = 1.84). All the previously mentioned municipalities border with *São Paulo* Municipality, such as *Barueri and São Caetano do Sul*, with the exception of *Igaratá* Municipality, which is located nearly 100 kilometers away from *São Paulo* by road.

## Discussion

In this paper, we adapted the methodology used by Desjardins et al. (2020), using three periods to detected emerging clusters of COVID-19 in São Paulo State, Brazil, utilizing confirmed case data by the Brazil IO (Brasil IO, 2020). It is important to highlight that the relative risk throughout the *São Paulo* State increased during the three periods analyzed. In the first period (February 25^th^ – March 24^th^, 2020) within 28 days from the first recorded case, we detected only five municipalities with a RR>1 and in the third period (February 25^th^ – May 5^th^, 2020), within 70 days from the first recorded case, we detected 23 municipalities with a RR>1. These results show that within our study period, there are 4.6 times more municipalities with more observed than expected cases for *São Paulo* State. Our results show that São Paulo Municipality is a hotspot of COVID-19 and this is highlighted by the location of the clusters with a RR>1 in the second and third periods. However, after 70 days of virus dispersion in the State of São Paulo, we observed municipalities with a RR>1 in almost all of the emerging clusters.

In the first period analyzed (February 25^th^ – March 24^th^, 2020), twenty municipalities had level of relative risk of COVID-19, but no clusters were detected; while all municipalities with reported COVID-19 incidence have a population greater than 50,000 inhabitants. In the second period (February 25^th^ - April 15^th^, 2020) and third period (February 25^th^ - May 5^th^, 2020), clusters with a RR> 1 were located around the municipality of São Paulo. However, in the second period, 25 municipalities with a population less than 50,000 inhabitants started to exhibit COVID-19 incidence. In the third period, the number of municipalities with reported COVID-19 incidence increased to 117, showing the spread to the countryside of the State. This reinforces a recent discovery by the Fundação Instituto Oswaldo Cruz (FIOCRUZ), where COVID-19 is arriving rapidly in medium-sized municipalities (20 to 50 thousand inhabitants), tending to generate the growth of transmission cycles for small cities. This is worrisome because health services are not prepared for this demand, leading to their burden on urban centers of reference in São Paulo State (FIOCRUZ, 2020).

Despite the contributions of our study, there are a few limitations that are worth pointing out. First, the cylindrical shapes of the cluster detection are likely not the true shape of the COVID-19 clusters. Second, we utilized lab confirmed case data, which does not capture the true burden of the epidemic in São Paulo State. We encourage researchers to examine our results and future work can understand the transmission dynamics of COVID-19 in the State and identify predictors of the epidemic. Third, future work can adjust for significant predictors of COVID-19 when computing the space-time scan statistic Finally, we used a prospective space-time statistical model, so we disregard clusters that “disappeared” during the study period and were not active on May 5^th^, 2020.

## Conclusions

This research presented an analysis of the dynamics of the expansion of COVID-19 based on the number of daily cases available in Brasil.IO’s database, with the intent of identifying emerging space-time clusters active in the municipalities of the State of São Paulo in three different time periods. We detected three significant active clusters in the State of *São Paulo* on May 5, 2020. Therefore, this space-time approach to map emerging clusters will allow decision-makers to identify statistically significant hotspots of COVID-19 cases. Regional Health Departments (DRS) of the State of São Paulo are responsible for coordinating the activities in regional level. These departments can use the results to optimize intersectoral coordination and organization of health care needs, specifically in relation to bed availability and pulmonary mechanical ventilator. In turn, this can improve the management of resources in the State of São Paulo. These results coupled with stay-home-orders orders (social confinement) can inform authorities on the impact of such measures to limit the propagation of COVID19 cases. As such, our results can support restrictive measures of social distancing that can be taken to minimize viral transmission.

## Data Availability

The COVID-19 case data is available at https://brasil.io/dataset/covid19/

